# How many are at increased risk of severe COVID-19 disease? Rapid global, regional and national estimates for 2020

**DOI:** 10.1101/2020.04.18.20064774

**Authors:** Andrew Clark, Mark Jit, Charlotte Warren-Gash, Bruce Guthrie, Harry HX Wang, Stewart W Mercer, Colin Sanderson, Martin McKee, Christopher Troeger, Kanyin I Ong, Francesco Checchi, Pablo Perel, Sarah Joseph, Hamish P Gibbs, Amitava Banerjee, CMMID COVID-19 working group, Rosalind M Eggo

## Abstract

**Background:** The risk of severe COVID-19 disease is known to be higher in older individuals and those with underlying health conditions. Understanding the number of individuals at increased risk of severe COVID-19 illness, and how this varies between countries may inform the design of possible strategies to shield those at highest risk.

**Methods:** We estimated the number of individuals at increased risk of severe COVID-19 disease by age (5-year age groups), sex and country (n=188) based on prevalence data from the Global Burden of Disease (GBD) study for 2017 and United Nations population estimates for 2020. We also calculated the number of individuals without an underlying condition that could be considered at-risk because of their age, using thresholds from 50-70 years. The list of underlying conditions relevant to COVID-19 disease was determined by mapping conditions listed in GBD to the guidelines published by WHO and public health agencies in the UK and US. We analysed data from two large multimorbidity studies to determine appropriate adjustment factors for clustering and multimorbidity.

**Results:** We estimate that 1.7 (1.0 - 2.4) billion individuals (22% [15-28%] of the global population) are at increased risk of severe COVID-19 disease. The share of the population at increased risk ranges from 16% in Africa to 31% in Europe. Chronic kidney disease (CKD), cardiovascular disease (CVD), diabetes and chronic respiratory disease (CRD) were the most prevalent conditions in males and females aged 50+ years. African countries with a high prevalence of HIV/AIDS and Island countries with a high prevalence of diabetes, also had a high share of the population at increased risk. The prevalence of multimorbidity (>1 underlying conditions) was three times higher in Europe than in Africa (10% vs 3%).

**Conclusion:** Based on current guidelines and prevalence data from GBD, we estimate that one in five individuals worldwide has a condition that is on the list of those at increased risk of severe COVID-19 disease. However, for many of these individuals the underlying condition will be undiagnosed or not severe enough to be captured in health systems, and in some cases the increase in risk may be quite modest. There is an urgent need for robust analyses of the risks associated with different underlying conditions so that countries can identify the highest risk groups and develop targeted shielding policies to mitigate the effects of the COVID-19 pandemic.

**Research in context:** *Evidence before this study:* As the COVID-19 pandemic evolves, countries are considering policies of ‘shielding’ the most vulnerable, but there is currently very limited evidence on the number of individuals that might need to be shielded. Guidelines on who is currently believed to be at increased risk of severe COVID-19 illness have been published online by the WHO and public health agencies in the UK and US. We searched PubMed (“Risk factors” AND “COVID-19”) without language restrictions, from database inception until April 5, 2020, and identified 62 studies published between Feb 15, 2020 and March 20, 2020. Evidence from China, Italy and the USA indicates that older individuals, males and those with underlying conditions, such as CVD, diabetes and CRD, are at greater risk of severe COVID-19 illness and death.

*Added value of this study:* This study combines evidence from large international databases and new analysis of large multimorbidity studies to inform policymakers about the number of individuals that may be at increased risk of severe COVID-19 illness in different countries. We developed a tool for rapid assessments of the number and percentage of country populations that would need to be targeted under different shielding policies.

*Implications of all the available evidence:* Quantifying how many and who is at increased risk of severe COVID-19 illness is critical to help countries design more effective interventions to protect vulnerable individuals and reduce pressure on health systems. This information can also inform a broader assessment of the health, social and economic implications of shielding various groups.

## Introduction

Emerging evidence from China, Europe and the USA has shown a consistently higher risk of severe COVID-19 disease in older individuals and those with underlying health conditions.^1-3^ Mild and moderate cases are those with symptoms but without evidence of pneumonia or dyspnea, and low mortality is expected in this group. Severe cases have those symptoms, would require hospitalisation if available, and are at risk of increased mortality.^4,5^ In a recent report from the USA, underlying conditions were reported in 71% (732/1037) of individuals hospitalised with COVID-19 and 94% (173/184) of deaths.^1^ The World Health Organization (WHO) as well as public health agencies in countries including the UK and US, have issued guidelines on who is considered to be at increased risk of severe COVID-19 illness.^6-8^ This includes individuals with cardiovascular disease (CVD), chronic kidney disease (CKD), diabetes, chronic respiratory disease (CRD) and a range of other chronic conditions. Such conditions increase the risk of needing hospital-based treatment such as oxygen supplementation or mechanical ventilation. A large proportion of the additional health care burden of COVID-19 epidemics is likely to result from infection of those with underlying conditions.

Identifying at-risk populations is important not only for making projections of the likely health burden in countries,^9,10^ but also for the design of effective strategies that aim to reduce the risk of transmission to people in target groups. This is sometimes termed shielding, defined as “a measure to protect extremely vulnerable people by minimising interaction between those who are extremely vulnerable and others.”^11^ It has the potential to reduce mortality in vulnerable groups (direct benefits), while at the same time mitigating the expected surge in demand for hospital beds (indirect benefits), which is critical where systems risk being strained by the national epidemic. However, trying to shield an excessive proportion of a population may strain country resources and reduce the overall effectiveness of shielding. A detailed assessment of the number of at-risk individuals can inform possible shielding strategies. If a vaccine becomes available in the future, it could also be used to inform the process of prioritising different groups, based on risk.

The aim of this work is to provide global, regional and national estimates of the numbers of individuals at increased risk of severe COVID-19 disease by virtue of their underlying medical conditions during 2020. The risk of severe COVID-19 disease is not binary, but unless tiered approaches are adopted, the criterion for whom to shield is, so our estimates seek to capture all those ‘at increased risk’ (UK terminology) or ‘at higher risk’ (US terminology) based on current guidelines.

## Methods

### Prevalence of underlying health conditions

We used the list of conditions thought to increase the risk of severe COVID-19 illness, based on current guidelines from WHO and public health agencies in the UK and US (supplementary appendix)^6-8^ and mapped these to eleven categories of underlying health conditions in the Global Burden of Disease Study (GBD). The mapping was completed by a clinical epidemiologist (CWG). Prevalence estimates were extracted for the following disease categories by age, sex and country: (1) CVD, including CVD caused by hypertension; (2) CKD, including CKD caused by hypertension; (3) CRD; (4) chronic liver disease; (5) diabetes; (6) cancers with direct immunosuppression; (7) cancers without direct immunosuppression, but with possible immunosuppression caused by treatment; (8) HIV/AIDS; (9) tuberculosis; (10) chronic neurological disorders; and (11) sickle cell disorders.

We estimate the current number of individuals with underlying conditions making them at risk of severe COVID-19 disease by age (5-year age groups), sex and country for 188 countries. Data on the prevalence of underlying conditions were extracted by age, sex and country from the GBD study for 2017^12^ and combined with United Nations mid-year population estimates for 2020.^13^ For this analysis, older individuals without underlying conditions were not considered to be at increased risk.

Asthma is relatively common, and mild asthma is not listed as a high risk group for shielding so we modified GBD estimates of asthma to account only for moderate-to-severe cases (defined as British Thoracic Society Steps 4, 5 and 6).^14^ Based on published evidence from the UK we assumed these were 15% of total asthma cases aged <5 years, 17% aged 5-19 years, 23% aged 20-54 years, and 43% in those aged 55+ years.^15^

For HIV/AIDS, we included all populations, including those on ART. However, we carried out a sensitivity analysis to determine how estimates would change if we removed individuals using anti-retroviral therapy (ART) for instance if there is found to be no additional risk of HIV in individuals on ART. We used WHO national estimates for ART coverage among those living with HIV/AIDS.^16^

### Proportion with at least one underlying condition relevant to severe COVID-19 disease

The GBD study provides prevalence estimates for each disease category separately, but not for the prevalence of people in at least 1 of these categories. One problem was that diseases may cluster, for example if they are causally related, and we incorporated this effect for each country, sex and age group (but we omit subscripts for clarity). We first calculated *e*, which is the expected proportion of individuals with at least one condition assuming that the prevalences of the underlying conditions are independent (e.g. the fact that someone has diabetes does not affect their risk of getting cancer) as 1 minus the probability of not having any of the conditions c1, c2, c3….i.e. 1 – (1 – p_c1) × (1 – p_c2) × (1 – p_c3)…. We then estimated the proportion *P*, who have at least one underlying condition as *P = e* × *r*, where *r* is the ratio between the observed and expected percentage of individuals with at least one condition. The ratio *r* is drawn from evidence from two large cross-sectional multimorbidity studies in Scotland^17^ and Southern China^18^ (supplementary appendix).

### Adjustment for multimorbidity

In addition to providing estimates for *r*, the studies in Scotland and Southern China were also used to calculate the multimorbidity fraction i.e. the proportion of individuals with multiple (two or more) underlying conditions among those with at least one, by age group and sex. All analyses were done using disease categories that matched as closely as possible to the COVID-19-relevant categories defined in our analysis. In both studies this included: CVD (defined as the presence of one or more of coronary heart disease, hypertension, cerebrovascular disease, peripheral arterial disease, heart failure, or atrial fibrillation); chronic neurological disease (defined as one or more of dementia, multiple sclerosis and Parkinson’s disease); and CRD (defined as one or both of chronic obstructive pulmonary disease and bronchiectasis). Other COVID-related conditions listed above were counted separately. The GBD provide separate estimates for hypertensive heart disease and CKD due to hypertension, but it was not possible to make this distinction in the multimorbidity datasets, so all hypertension was included in the CVD category.

Using the data from both studies, we calculated pooled estimates of the ratio *r*, and the multimorbidity fraction by age and sex (supplementary appendix) and extrapolated these pooled estimates to all countries included in the analysis.

### Inclusion of older individuals without underlying conditions

Some countries have also considered older age as proxy for frailty and increased risk of severe COVID-19 illness. Although frailty correlates much more closely with mortality than chronological age, there is a well-established non-linear association between increasing age and frailty.^19^ We therefore calculated the number of individuals without an underlying condition that could be considered at-risk because of their age, using age thresholds ranging from 50-70 years. All age thresholds were evaluated in all regions. To calculate the total number at increased risk for different age thresholds, we added the number of older individuals without underlying conditions to our earlier estimates of the number of individuals with at least one underlying condition.

### Uncertainty

We generated low and high estimates using the lower and upper 95% confidence limits for the country population size, individual disease prevalences and age-specific multimorbidity fraction. We also varied *r*, the ratio between the observed and expected percentage of individuals with at least one condition, by a range informed by the multimorbidity studies.

All analyses are provided in an Excel spreadsheet, available at https://cmmid.github.io/topics/covid19/.

## Results

### Individuals with at least one underlying condition at risk of severe COVID-19 disease

We estimate that 1.7 billion (1.0 - 2.4) individuals (22% [15-28%] of the global population) have at least one underlying condition that could increase their risk of severe COVID-19 disease (table 1, supplementary appendix). This value does not include older individuals without underlying conditions. The prevalence of one or more condition was approximately 10% by age 25 years, 33% by 50 years, and 66% by 70 years, and similar for males and females (figure 1). The most prevalent conditions in those aged 50+ years were CKD, CVD, CRD and diabetes.

**Table 1.** Number of individuals in millions (% of total population) at increased risk of severe COVID-19 illness by age, number of conditions, region and age threshold.

|  | Africa | Asia | Europe | Latin America and the Caribbean | Northern America | Oceania | Global |
| --- | --- | --- | --- | --- | --- | --- | --- |
| **No conditions** |  |  |  |  |  |  |  |
| <15 years | 519 (39) | 1042 (22) | 117 (16) | 152 (23) | 65 (18) | 10 (23) | **1905 (24)** |
| 15-49 years | 534 (40) | 1986 (43) | 274 (37) | 292 (45) | 147 (40) | 17 (40) | **3249 (42)** |
| 50-54 years | 25 (2) | 184 (4) | 33 (4) | 23 (4) | 14 (4) | 2 (4) | **281 (4)** |
| 55-59 years | 18 (1) | 137 (3) | 29 (4) | 18 (3) | 13 (3) | 1 (3) | **216 (3)** |
| 60-64 years | 12 (1) | 96 (2) | 23 (3) | 13 (2) | 10 (3) | 1 (2) | **154 (2)** |
| 65-69 years | 7 (1) | 69 (1) | 16 (2) | 8 (1) | 7 (2) | 1 (2) | **108 (1)** |
| 70+ years | 7 (0) | 74 (2) | 23 (3) | 10 (2) | 9 (2) | 1 (2) | **123 (2)** |
| All ages | 1121 (84) | 3588 (77) | 516 (69) | 515 (79) | 264 (72) | 32 (76) | **6036 (78)** |
| **One condition only** |  |  |  |  |  |  |  |
| <15 years | 20 (1) | 44 (1) | 3 (0) | 4 (1) | 2 (0) | 0 (1) | **72 (1)** |
| 15-49 years | 101 (8) | 369 (8) | 49 (7) | 45 (7) | 20 (5) | 3 (7) | **588 (8)** |
| 50-54 years | 14 (1) | 83 (2) | 14 (2) | 10 (2) | 7 (2) | 1 (1) | **128 (2)** |
| 55-59 years | 12 (1) | 78 (2) | 17 (2) | 10 (2) | 8 (2) | 1 (2) | **127 (2)** |
| 60-64 years | 11 (1) | 69 (1) | 18 (2) | 10 (1) | 9 (3) | 1 (2) | **118 (2)** |
| 65-69 years | 8 (1) | 62 (1) | 18 (2) | 8 (1) | 9 (2) | 1 (2) | **105 (1)** |
| 70+ years | 11 (1) | 93 (2) | 40 (5) | 15 (2) | 17 (5) | 1 (3) | **178 (2)** |
| All ages | 178 (13) | 797 (17) | 159 (21) | 103 (16) | 72 (20) | 7 (17) | **1316 (17)** |
| **Multiple (two or more) conditions** |  |  |  |  |  |  |  |
| <15 years | 1 (0) | 3 (0) | 0 (0) | 0 (0) | 0 (0) | 0 (0) | **4 (0)** |
| 15-49 years | 13 (1) | 53 (1) | 8 (1) | 7 (1) | 3 (1) | 0 (1) | **85 (1)** |
| 50-54 years | 4 (0) | 23 (0) | 4 (1) | 3 (0) | 2 (0) | 0 (0) | **35 (0)** |
| 55-59 years | 4 (0) | 27 (1) | 6 (1) | 4 (1) | 3 (1) | 0 (1) | **44 (1)** |
| 60-64 years | 4 (0) | 29 (1) | 8 (1) | 4 (1) | 4 (1) | 0 (1) | **50 (1)** |
| 65-69 years | 4 (0) | 33 (1) | 9 (1) | 4 (1) | 5 (1) | 0 (1) | **56 (1)** |
| 70+ years | 9 (1) | 80 (2) | 37 (5) | 13 (2) | 16 (4) | 1 (3) | **156 (2)** |
| All ages | 41 (3) | 247 (5) | 72 (10) | 35 (5) | 32 (9) | 3 (7) | **430 (6)** |
| **At increased risk of severe COVID-19 disease** |  |  |  |  |  |  |  |
| At least one condition | 218 (16) | 1044 (23) | 231 (31) | 137 (21) | 104 (28) | 10 (24) | **1746 (22)** |
| ...plus age 50+ years | 286 (21) | 1605 (35) | 356 (48) | 209 (32) | 157 (43) | 16 (37) | **2628 (34)** |
| ...plus age 55+ years | 261 (20) | 1420 (31) | 322 (43) | 186 (29) | 143 (39) | 14 (34) | **2346 (30)** |
| ...plus age 60+ years | 244 (18) | 1284 (28) | 293 (39) | 168 (26) | 130 (35) | 13 (30) | **2130 (27)** |
| ...plus age 65+ years | 238 (18) | 1199 (26) | 288 (39) | 160 (25) | 131 (35) | 12 (29) | **2028 (26)** |
| ...plus age 70+ years | 230 (17) | 1137 (25) | 271 (36) | 152 (23) | 122 (33) | 12 (28) | **1923 (25)** |

**Figure 1.**
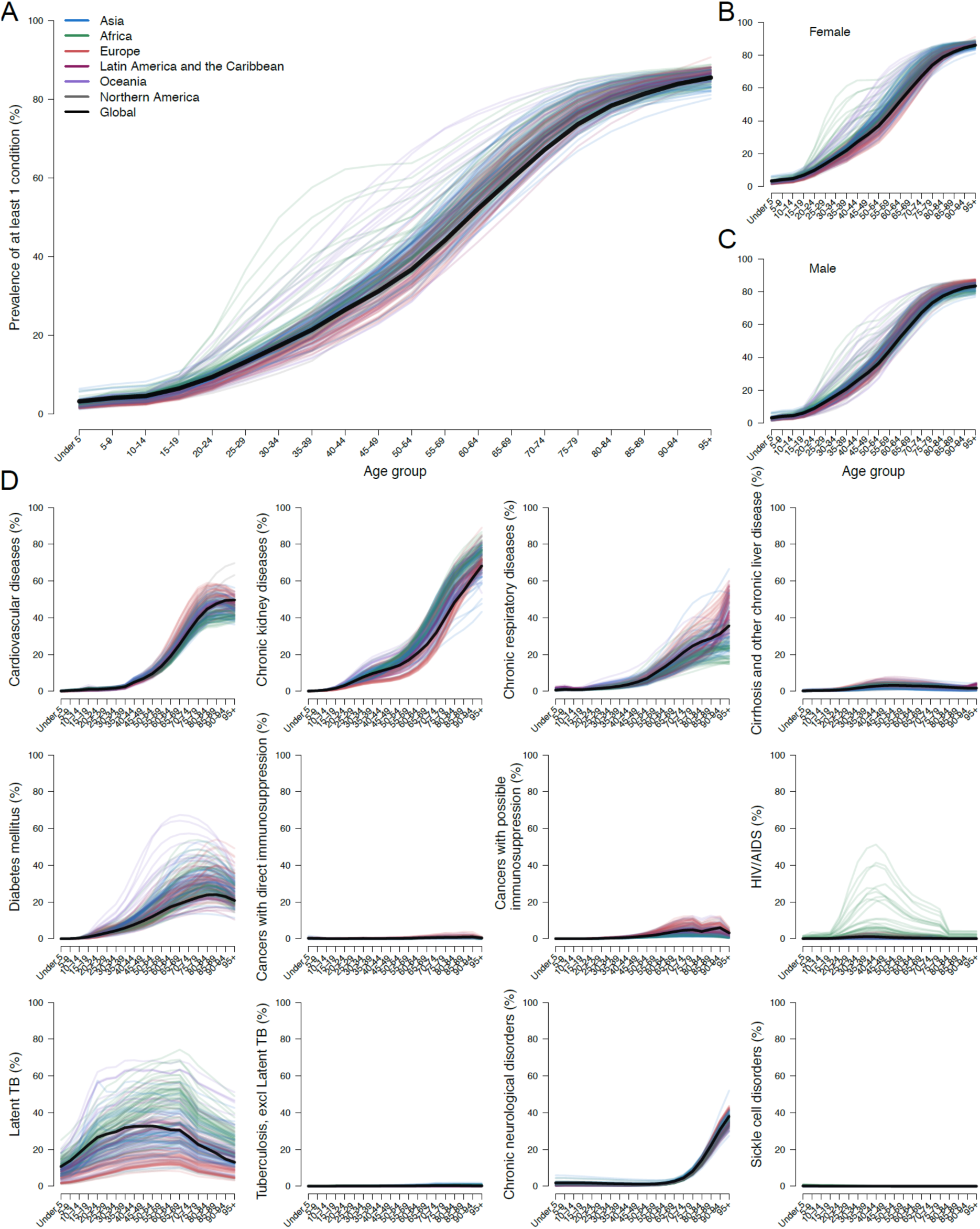
Proportion of individuals with at least one underlying condition by age, sex, country and region; and prevalence of each underlying condition by age and region. We excluded latent tuberculosis from our analysis but include it here to show the extent of overall TB that was excluded. Only the global estimates (solid black line) and country lines are shown (thin lines), but thin lines are coloured to indicate the UN population region they belong to. For example, some African countries (green) have a high HIV prevalence, and some Island countries in Oceania (purple) have a high prevalence of diabetes.

Based on crude proportions without age-standardisation, the share of the population at-risk ranged from 16% in Africa to 31% in Europe (table 1)(figures 2 and 3). The highest estimates were in European and other high-income countries with a high prevalence of CKD, CVD, diabetes and CRD in older ages. African countries with a high prevalence of HIV/AIDS and Island countries with a high prevalence of diabetes, also had a high share of the population at increased risk.

**Figure 2.**
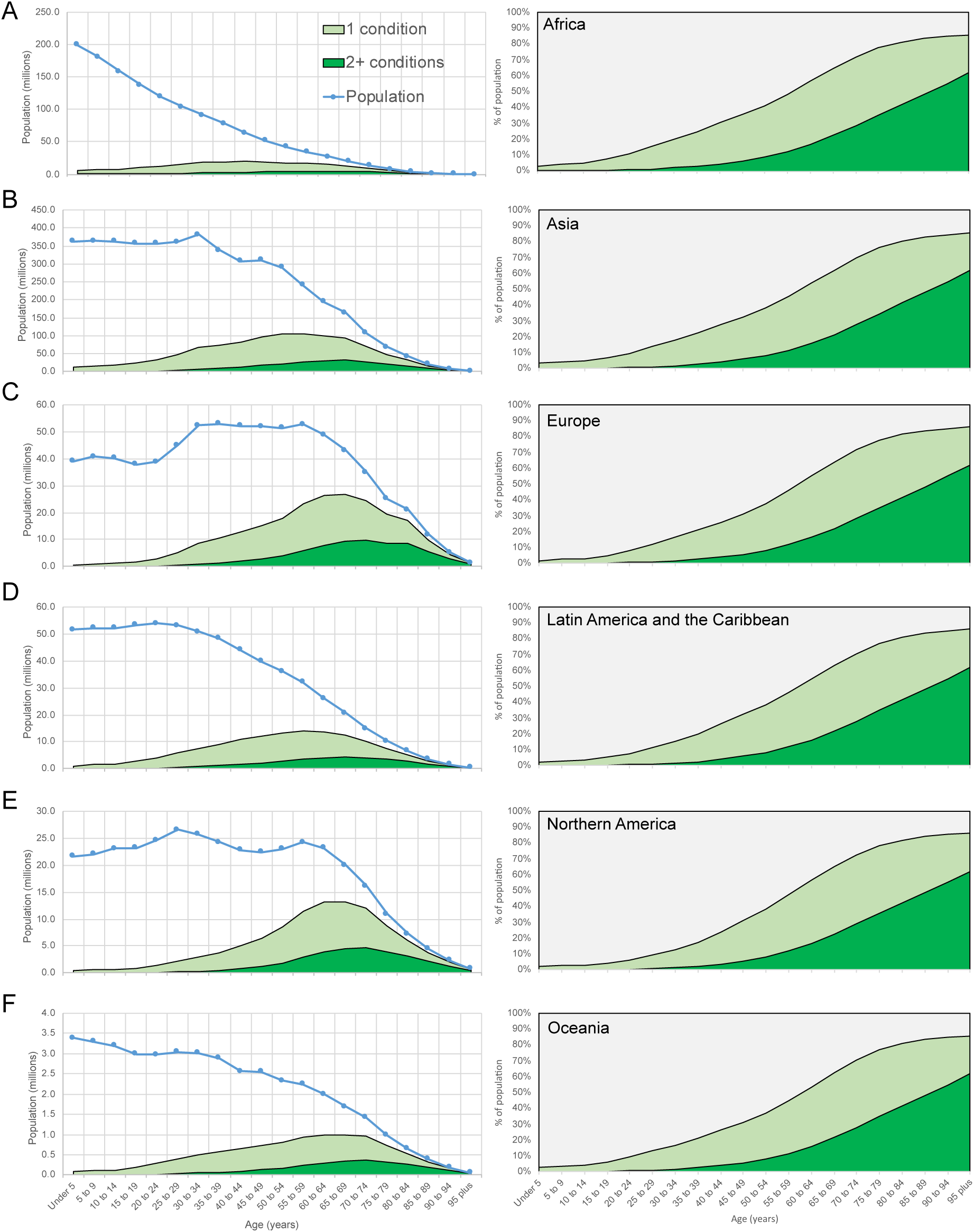
Population and number of individuals with 1 and 2+ conditions by age and region (left); prevalence of 1 and 2+ conditions by age and region (right) Panels A-F represent each UN population region. The plots on the left side show the population by age (blue line), those with only 1 condition (light green area) and those with multiple (two or more) conditions (dark green). The plots on the right show prevalence of one and multiple conditions by age.

**Figure 3.**
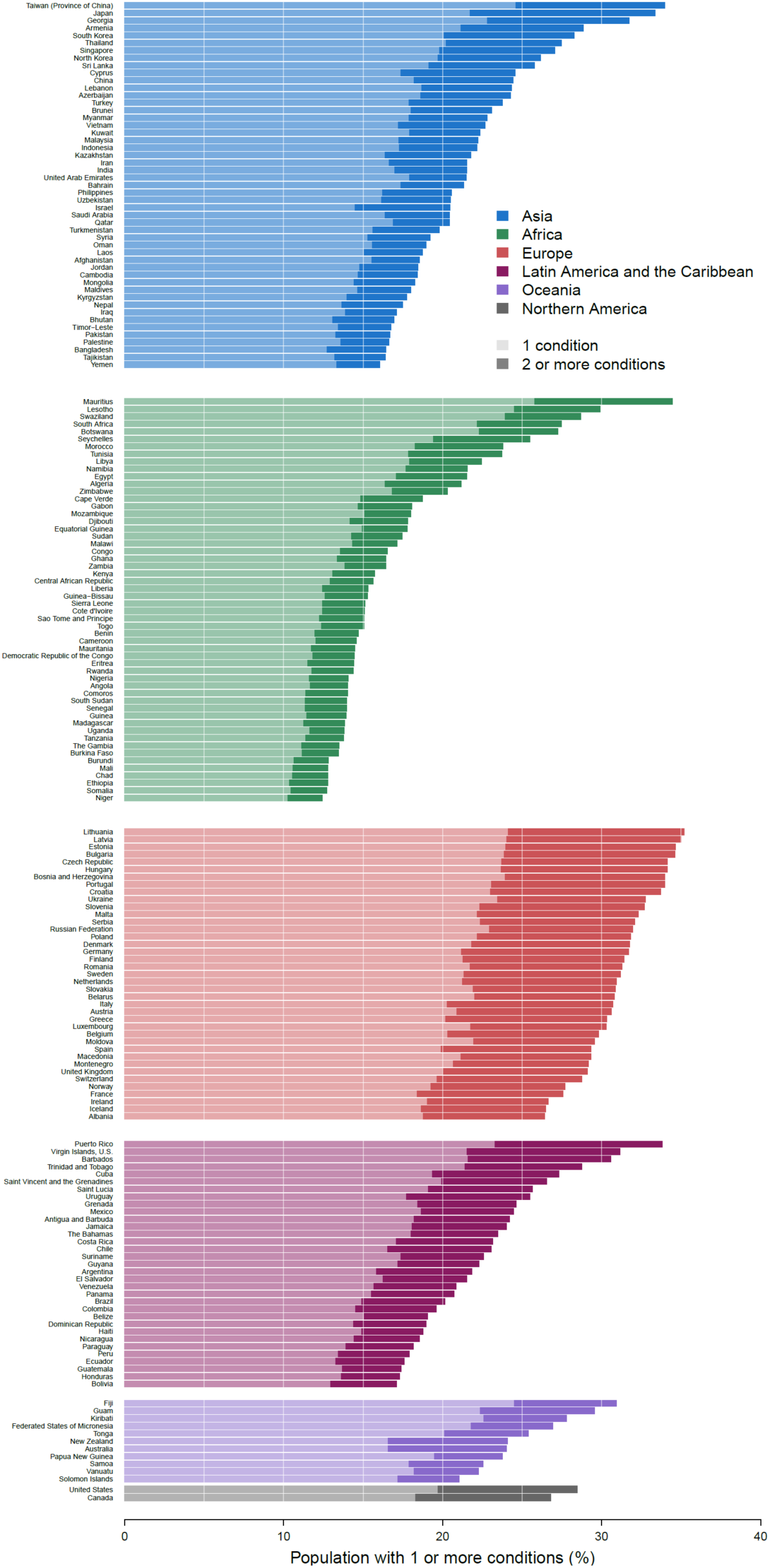
Proportion of population with underlying conditions relevant to COVID-19 disease by number of conditions, country and region. Each country is given, grouped by UN population regions. The lighter bars give the percentage of the population with 1 underlying condition, and darker the percentage of the population with more than 1 underlying condition.

In African countries with high HIV prevalence excluding those on ART reduced the at-risk proportion e.g. from 27 to 20% in Botswana, 30 to 24% in Lesotho, 27 to 23% in South Africa and 29 to 19% in Swaziland.

We estimate that 23% (range 15-38%) of the working age population (15-64 years) have at least one underlying condition (figure 4). This proportion was particularly high in some European countries, and Island countries with high rates of diabetes (figure 3). CKD and diabetes were the most common conditions in this age range.

**Figure 4.**
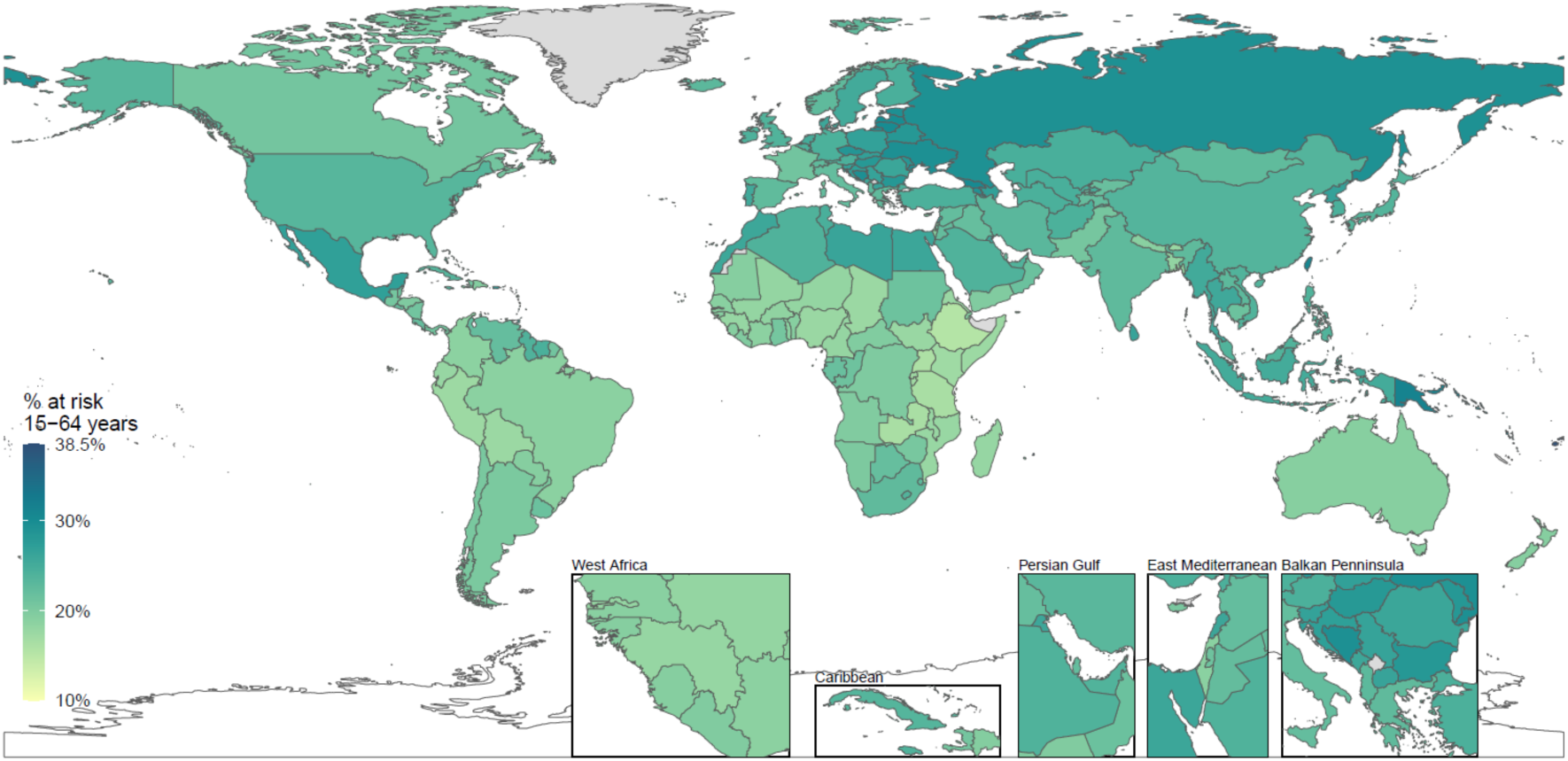
Proportion of working age population (15-64 years) with at least one underlying condition relevant to COVID-19. Countries shaded in darker green have a higher proportion of their working age population with at least one underlying condition relevant to COVID-19. Lighter shaded countries have a lower proportion of their working age population with at least one underlying condition.

### Individuals with multimorbidities associated with severe COVID-19 disease

Among the 1.7 billion individuals estimated to be at increased risk, we estimate that 0.4 billion (0.2 – 0.7) individuals (6% [3-8%] of the global population) are living with two or more conditions relevant to COVID-19 outcomes in the year 2020 (table 1, supplementary appendix). As expected, this proportion was higher in regions with an older age profile. The prevalence of multimorbidity (two or more underlying conditions) was three times higher in Europe than in Africa (10% vs 3%) (table 1).

### Inclusion of older individuals without underlying conditions

The number of individuals without an underlying condition that could be considered at-risk solely because of their age varied by region and choice of age threshold (table 1). In Africa, around 5% of individuals aged 50+ years are estimated to have no underlying conditions linked to severe COVID-19 disease. A similar proportion is estimated at age 65+ years in Europe, North America and Oceania, and at 60+ years in Asia and Latin America.

### New analysis of cross-sectional multimorbidity studies

The ratio *r* was broadly consistent by age, sex and study (supplementary appendix). When extrapolating this value to other countries, we used a transparent ratio of 0.9 for all age groups and varied this between 0.7 and 1.0 in our low and high estimates. The resulting national estimates of *P* were constrained to be no less than each country’s single most prevalent condition.

We estimated that <10% of individuals with underlying conditions have multiple conditions before 30 years of age, but this increased to about 40% by age 70 years (supplementary appendix). The most common combinations in both studies were CVD plus diabetes, CVD plus CKD and CVD plus CRD.

## Discussion

Based on current guidelines and prevalence estimates from GBD, we calculate that one in every five individuals worldwide has a condition that is on the list of those at increased risk of severe COVID-19 disease. However, for many of these individuals the underlying condition will be undiagnosed or not severe enough to be captured in health systems. Indeed, most will not progress to severe COVID-19 disease or death if infected with SARS-Cov-2.^1^ However, because severe COVID-19 outcomes are disproportionately found in individuals with underlying conditions,^1^ protecting these groups may be one of the most effective ways to reduce mortality and demand for hospital beds.^20^

Our estimates of the numbers at-risk are based on prevalence estimates extracted from the GBD. The GBD study produces estimates of disease prevalence for all ages, both sexes, and for 195 territories and countries from 1990-2017.^21^ Because the GBD produces internally comparable estimates for a comprehensive list of diseases, these estimates are well suited to compare prevalence of disease across locations.

A recent analysis from Sweden^22^ provides an opportunity to evaluate our method, and compare GBD prevalence estimates to those derived from electronic health record (EHR) data. When we applied our method, based on the calculation of *e* and *r*, to the prevalence of each condition reported in the study, we were able to reproduce the same percentage share of the population at increased risk. This provides some reassurance that our method will provide a reasonable approximation of the at-risk proportion if the same input data on disease prevalence is used. The GBD prevalence estimates for Sweden were substantially higher than the estimates from the EHR data, and so we estimate a much higher proportion of the population to be at increased risk (35 vs 22%). Many of the prevalent cases estimated by GBD would not have been severe enough to be recorded in EHRs, which is likely the major reason for the difference. For example, over half of the CKD cases included in GBD prevalence estimates represent early stage disease (CKD stage 1 or 2) which is common and rarely has symptoms.^23^ Several other underlying conditions estimated by GBD are also likely to be undiagnosed in many individuals e.g. hypertensive heart disease, compensated chronic liver disease, early onset diabetes etc. This has important implications for the interpretation of our estimates because it suggests, based on this initial comparison exercise for one country, that around one-third of those estimated to have an underlying condition may be undiagnosed or not severe enough to be captured in EHRs. For these individuals, shielding policies may not identify them, and perhaps more importantly, their increase in risk may be quite modest.

If a safe and effective vaccine is produced, then our estimates provide an indication of the volumes that would be required for vaccination of at-risk individuals globally. In the absence of a vaccine, a key option to mitigate the pandemic is to shield at-risk individuals by more intensive physical-distancing measures than those in the wider population. This may be especially important at times and places where health systems risk being overwhelmed by cases. Other infection control measures include provision of personal protective equipment and intensive testing of health and social care workers in maximum contact with at-risk individuals. At a minimum, timely and effective communication should be provided to communities on who within them is at increased risk. Among those who are identified, governments will rely heavily on their adherence to guidelines. This would allow practical individual-level steps to be taken, such as increased hygiene, physical isolation and home-delivered food and medical care.^6^ As more evidence emerges on the risk associated with different conditions, guidelines could be refined and shielding policies tailored to different risk groups e.g. isolation for those at very high risk, and less stringent measures (e.g. social distancing) for those with a lower level of increased risk. Community-level shielding approaches, including vacating houses or public buildings to physically isolate small groups of people at increased risk, may also be considered, though these will require stringent infection control arrangements, especially in crowded settings,^24^ and adherence may be low if at-risk individuals are daily wage earners or people caring for children e.g. grandparents.^25^

The association between the prevalence of underlying conditions and other national characteristics, such as economic development, is complex. The prevalence of many of these conditions (HIV/AIDS may be an exception) reflect the epidemiological transition^26^ but survival with these conditions may reflect the performance of the health system.^27^ Hence, it is important to look at the data for each country, which goes beyond what we can report in this paper. We provide a spreadsheet tool (available at https://cmmid.github.io/topics/covid19/) that can be used for rapid assessment (and visualisation) of the estimated number and percentage of country populations targeted under different shielding policies. This allows different health conditions to be included/excluded, different age thresholds to be assessed, and different choices about key assumptions e.g. estimates of the ratio *r*, and the multimorbidity fraction by age. Prevalence data included in our spreadsheet can be updated as more evidence emerges on the importance (or otherwise) of specific conditions and their severities e.g. early stage CKD, simple hypertension etc.

Our analysis found a substantial number of at-risk individuals in the working age range. Strict shielding measures could cause considerable economic disruption to these individuals and their families and have a detrimental effect on the wider economy. For some at-risk individuals, particularly daily wage earners in low-income countries, it will be important to have alternative options to isolation. For example, there is growing evidence in support of face-masks as a means to prevent transmission by those wearing them.^28^ If proven to be effective, or other measures emerge,^29^ this could be a practical way of reducing exposure among those who are unable to avoid contact with others. Alternative shielding approaches, so that target groups are supported to reduce physical contacts (e.g. through incentives to reduce or abstain from work), can also be considered.

We based our analysis on underlying conditions listed in guidelines published by the WHO and public health agencies in the UK and US. As with seasonal influenza vaccination, these guidelines tend not to be exhaustive, and deliberately permit clinical judgement. As our understanding of this disease evolves, it should be possible to provide greater clarity on the risk of severe COVID-19 disease associated with different underlying conditions at different severities e.g. early stage CKD, compensated liver cirrhosis, moderately severe asthma etc.^6^ A detailed multivariate analysis of risk is urgently needed because nearly all underlying conditions are likely to be at least partly confounded by age. Knowledge of the mode of action of the virus is increasing rapidly, especially in relation to its action on cells with ACE2 receptors other than in the lung, in particular in the endothelium and pancreas, which the inflammatory processes associated with diabetes may be involved in cytokine storms that can be fatal.^30^

We estimated a similar number of males and females to be at increased risk, although early evidence from China, and later studies in other countries show a higher proportion of deaths in males.^31^ Earlier research in mice infected with SARS coronavirus also found an increased male susceptibility mediated by differences in oestrogen receptor signalling,^32^ while others have noted the concentration of genes involved in the immune system on the × chromosome.^33^ This is a clearly priority for further research.

We focused our analysis on the list of underlying chronic causes of disease available in GBD2017, and did not include risk factors, such as pregnancy, smoking, under-nutrition, obesity, working in the health and social care system, homelessness and general frailty e.g. individuals living in care homes and other facilities. The importance of these factors on the severity of COVID-19 is still to be determined, although some national guidelines already include frailty in decisions about treatment.^34^

We found that the age threshold for older people without underlying conditions led to a substantial increase in the overall number of people that may be considered at increased risk of severe COVID-19 disease. Individuals in developing countries may be frailer than those of an equivalent age in developed countries, and there may be other societal and economic considerations. Older individuals without these underlying conditions could suffer adverse mental health consequences from long periods of isolation. However, of the approximately 1 billion individuals aged 50+ years that we estimate to be at increased risk of severe COVID-19 disease (table 1) many may not be aware of their underlying condition.^35^ For example, CKD has a very high prevalence in many countries, but more than 90% of these cases are likely to be undiagnosed.^23^ Age thresholds could therefore play a critical role in shielding the large number of older individuals without a diagnosis. Indeed, while our analysis quantifies numbers who could benefit from shielding, in practice the low coverage of diagnosis and treatment for many chronic conditions in low-income settings means that the age threshold could largely determine the effective target group.

With the exception of HIV/AIDS our estimates do not account for the role of treatment or control of the underlying condition: for example, while some conditions are less prevalent in Africa than Europe, African patients may on average be at higher risk as they have less access to diagnosis and effective treatment. It is not yet known whether those with HIV are at increased risk of severe disease with COVID-19. Whilst it has been shown that widespread introduction of ART reduced the risk of hospitalisation and death associated with seasonal influenza,^36^ a substantial proportion of those on ART remain somewhat immunocompromised.^37,38^ Recent evidence from South Africa has shown that individuals living with HIV have an eight-fold higher risk of pneumonia hospitalisation associated with seasonal influenza, and a three-fold higher risk of pneumonia death.^39^ Until more evidence emerges, it may be necessary to include these individuals in shielding strategies, irrespective of ART status, with priority given to those not yet receiving treatment.^40^

We used data from two large studies to adjust for multimorbidity. Both studies could underestimate the prevalence of some conditions and therefore the extent of multimorbidity, although in Scotland, most of the included conditions were well recorded in routine health care, and in the Southern China study, underlying conditions were well communicated to patients, whose information was collected in a community household survey following a standard protocol. These studies cannot capture the global diversity of patterns of multimorbidity, which will differ in regions where, for example, there are high prevalences of HIV or sickle cell disorders. As multivariate analysis of the risk of serious COVID-19 become available, results that include combinations may provide more nuanced information to inform decisions about shielding.

Over the coming weeks and months, countries will need to determine mitigation strategies for the pandemic. Where feasible, shielding strategies could reduce the overall health burden by decreasing risk of infection to those most likely to experience severe disease and thus require health care. If implemented, shielding of at-risk individuals is likely to be required for several months. This may have a substantial impact on working-age people if they and their household contacts are less economically active for longer than the general population.

There is an urgent need for robust analyses of the risks associated with different underlying conditions so that countries can identify the highest risk groups and develop targeted shielding policies to mitigate the effects of the COVID-19 pandemic.

## Data Availability

Data used are publicly available on the Global Burden of Disease (2017) website, and the spreadsheet used for calculations is included.

## Acknowledgements

We acknowledge Jennifer Quint, Arminder Deol and Laurie Tomlinson for providing technical and clinical advice on specific diseases. We also acknowledge Ulla Griffiths and Palwasha Anwari for providing feedback on the spreadsheet.

The CMMID COVID-19 working group declare support from the following organisations: Bill and Melinda Gates Foundation (grants: OPP1183986, OPP1191821, INV-003174, OPP1180644, OPP1184344), RCUK/ESRC (grant: ES/P010873/1), UK Public Health Rapid Support Team, National Institute of Health Research (NIHR) Health Protection Research Unit (HPRU) in Modelling Methodology, European Commission (grant: 101003688), NIHR (grants: PR-OD-1017-20002, 16/137/109), NIHR EPIC grant (grant: 16/137/109), European Research Council Starting Grant (Action Numbers #757688, and #757699), Wellcome Trust (grants: 210758/Z/18/Z, 208812/Z/17/Z, 206250/Z/17/Z), Medical Research Council (MRC) London Intercollegiate Doctoral Training Program studentship (grants: MR/N013638/1), MRC (grant: MR/P014658/1), The Nakajima Foundation; The Alan Turing Institute, NIHR HPRU in Immunisation (grant: HPRU-2012-10096), Global Challenges Research Fund (GCRF) for the project “RECAP” managed through RCUK and ESRC (grant: ES/P010873/1) and Elrha’s Research for Health in Humanitarian Crises (R2HC) Programme. The R2HC programme is funded by the UK Government (DFID), the Wellcome Trust, and the UK NIHR.

## Supplementary appendix

**Figure 1.**
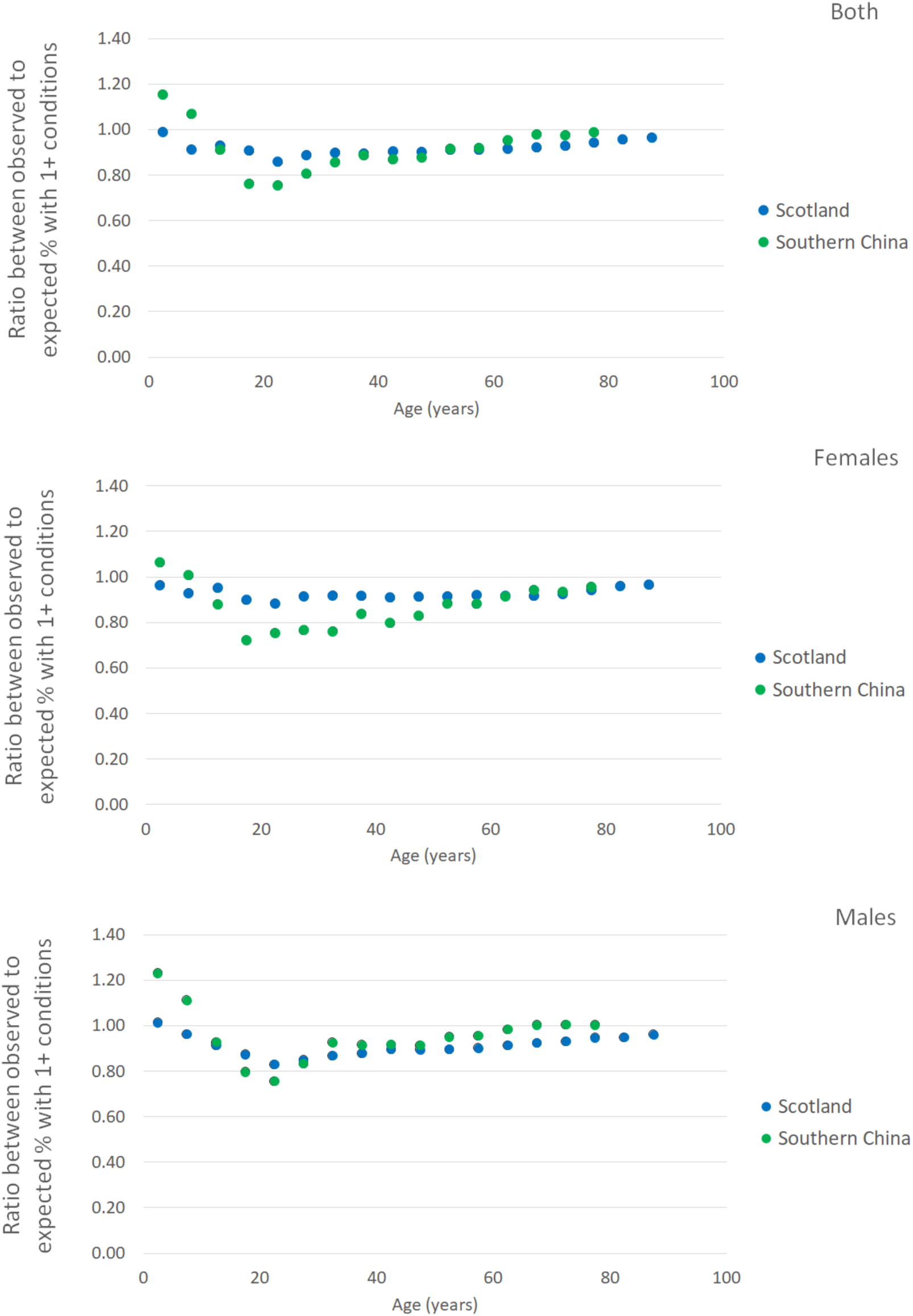
Ratio between observed and expected % of individuals with at least one condition by age from multimorbidity studies in Scotland and Southern China. For the analysis of both males and females combined, the mean of all age-specific values of r was 0.92 (range 0.86 to 0.99) in Scotland and 0.92 (range (0.75 – 1.15) in China. For the main analysis we assumed 0.9 across all ages (central estimates) for males and females combined in all countries and varied this between 0.7 (low estimates) and 1.0 (high estimates).

**Figure 2.**
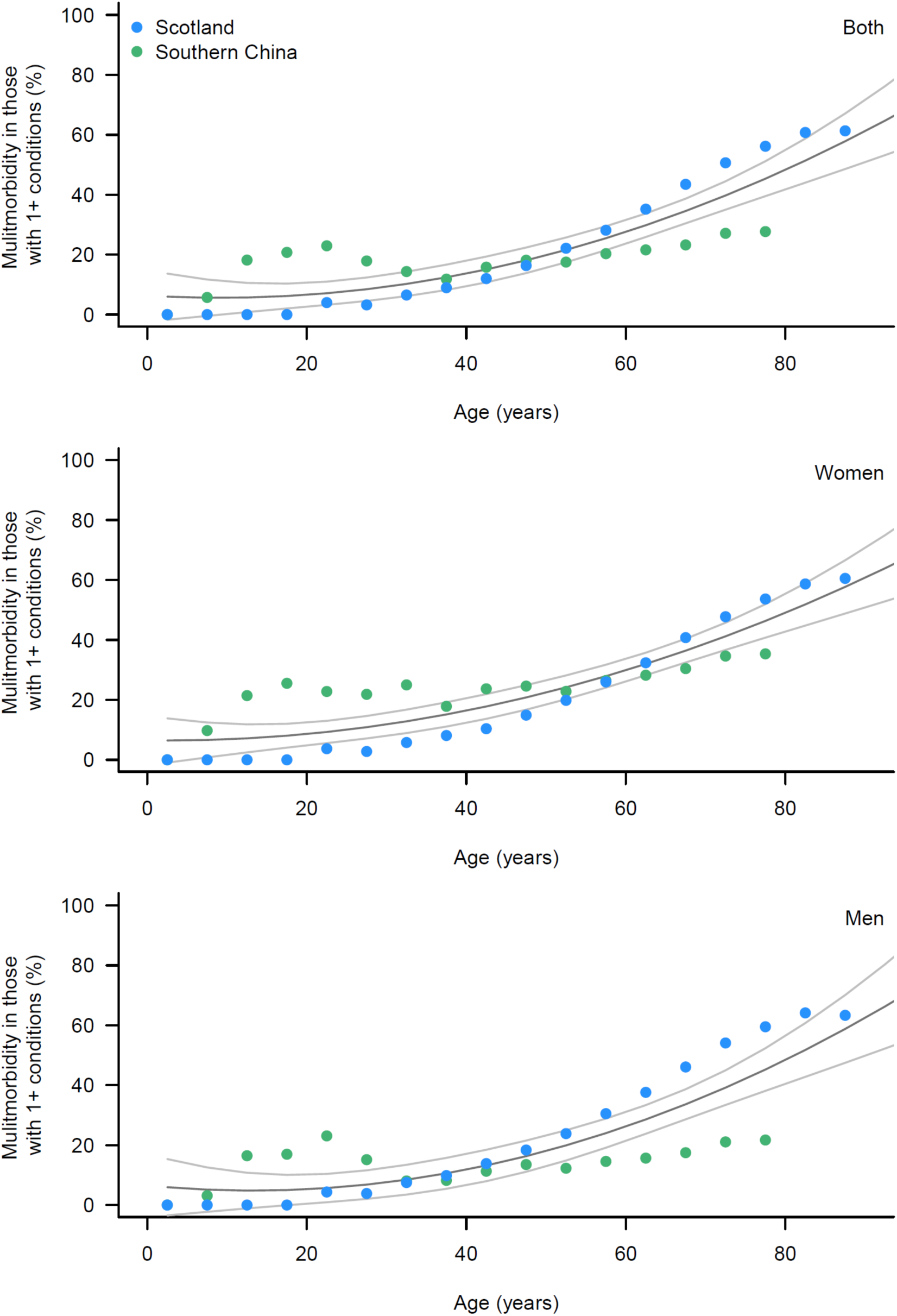
Multimorbidity fraction among those with at least one condition relevant for COVID-19 from cross-sectional studies in Scotland and Southern China. The grey lines represent pooled estimates and 95% confidence intervals based on a 2^nd^ order polynomial model fitted to all data points.

**Figure 3.**
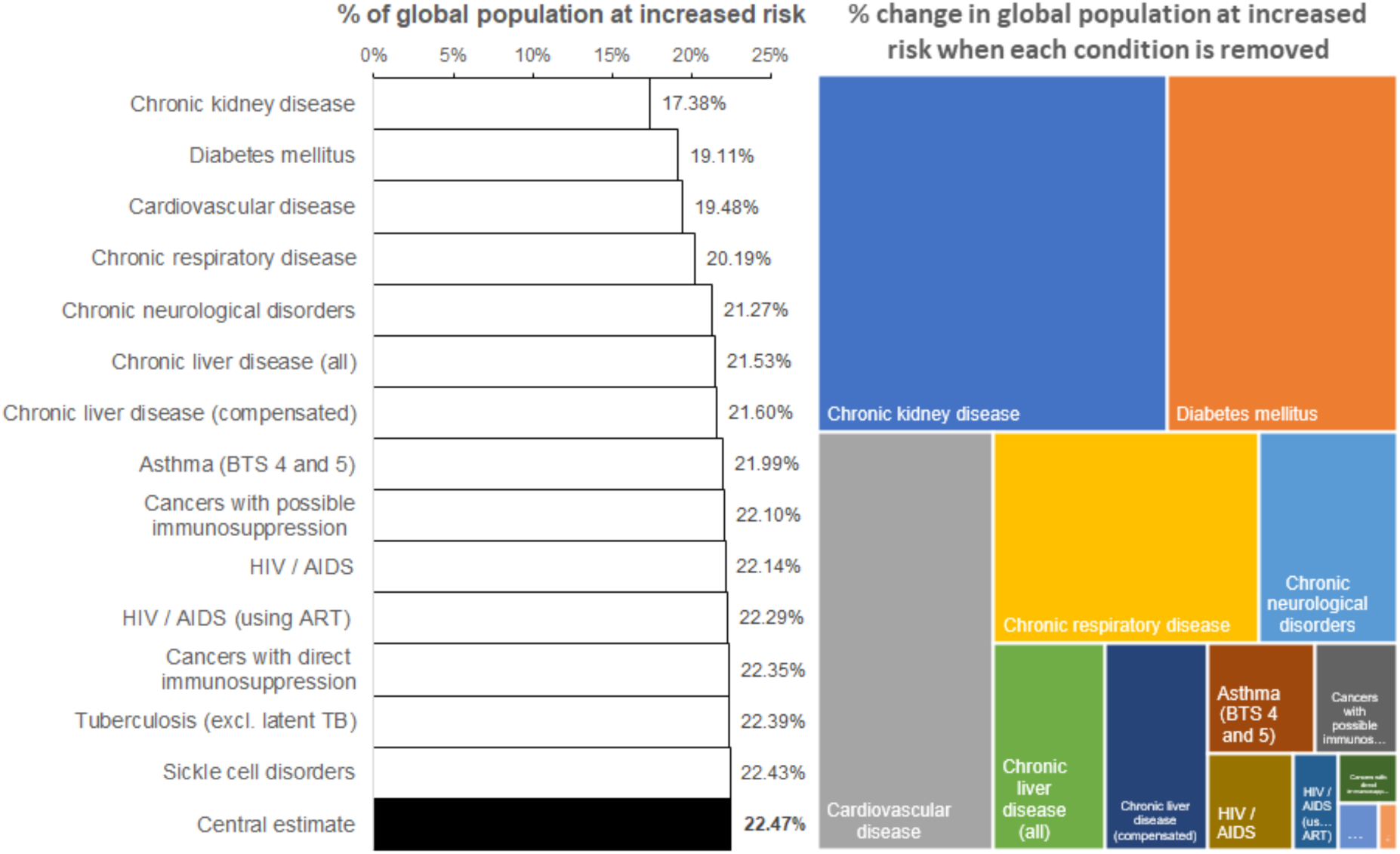
Percentage of global population at increased risk of severe COVID-19 illness (left panel) and influence on global estimates (right panel) when conditions are removed one at a time. The black shaded bar at the bottom represents the central estimate of the global population that are at increased risk. All other bars above show how this value changes when each of the conditions are removed one at a time. The most influential conditions are at the top of the bar chart and represent larger areas on the map shown on the right side.

**Table 1.** List of conditions included in GBD2017 with potential to increase the risk of severe COVID-19 illness.

| # | Category | Causes included in the Global Burden of Disease Study (GBD2017) |
| --- | --- | --- |
| 1 | HIV/AIDS | HIV/AIDS - Drug-susceptible Tuberculosis; HIV/AIDS - Multidrug-resistant Tuberculosis without extensive drug resistance; HIV/AIDS - Extensively drug-resistant Tuberculosis; HIV/AIDS resulting in other diseases |
| 2 | Tuberculosis* | Drug-susceptible tuberculosis; Multidrug-resistant tuberculosis without extensive drug resistance; Extensively drug-resistant tuberculosis |
| 3 | Cancers with direct immune suppression | Hodgkin lymphoma; Non-Hodgkin lymphoma; Multiple myeloma; Acute lymphoid leukemia; Chronic lymphoid leukemia; Acute myeloid leukemia; Chronic myeloid leukemia; Other leukemia; Other malignant neoplasms; Myelodysplastic, myeloproliferative, and other hematopoietic neoplasms |
| 4 | Cancers with possible immune suppression (from treatment therapy) | Lip and oral cavity cancer; Nasopharynx cancer; Other pharynx cancer; Esophageal cancer; Stomach cancer; Colon and rectum cancer; Liver cancer due to hepatitis B; Liver cancer due to hepatitis B; Liver cancer due to hepatitis B; Liver cancer due to hepatitis C; Liver cancer due to alcohol use; Liver cancer due to NASH; Liver cancer due to other causes; Gallbladder and biliary tract cancer; Pancreatic cancer; Larynx cancer; Tracheal, bronchus, and lung cancer; Malignant skin melanoma; Breast cancer; Cervical cancer; Uterine cancer; Ovarian cancer; Prostate cancer; Testicular cancer; Kidney cancer; Bladder cancer; Brain and nervous system cancer; Thyroid cancer; Mesothelioma |
| 5 | Cardio-vascular diseases | Rheumatic heart disease; Ischemic heart disease; Ischemic stroke; Intracerebral haemorrhage; Subarachnoid haemorrhage; Hypertensive heart disease; Non-rheumatic calcific aortic valve disease; Non-rheumatic degenerative mitral valve disease; Other non-rheumatic valve diseases; Myocarditis; Alcoholic cardiomyopathy; Other cardiomyopathy; Atrial fibrillation and flutter; Aortic aneurysm; Peripheral artery disease; Endocarditis; Other cardiovascular and circulatory diseases; Congenital heart anomalies |
| 6 | Chronic respiratory diseases | Chronic obstructive pulmonary disease; Silicosis; Asbestosis; Coal workers pneumoconiosis; Other pneumoconiosis; Asthma*; Interstitial lung disease and pulmonary sarcoidosis |
| 7 | Cirrhosis and other chronic liver diseases | Cirrhosis and other chronic liver diseases due to hepatitis B; Cirrhosis and other chronic liver diseases due to hepatitis C; Cirrhosis and other chronic liver diseases due to alcohol use; Cirrhosis and other chronic liver diseases due to other causes |
| 8 | Diabetes mellitus | Diabetes mellitus type 1; Diabetes mellitus type 2 |
| 9 | Chronic kidney diseases | Chronic kidney disease due to diabetes mellitus type 1; Chronic kidney disease due to diabetes mellitus type 2; Chronic kidney disease due to hypertension; Chronic kidney disease due to glomerulonephritis; Chronic kidney disease due to other and unspecified causes |
| 10 | Chronic neurological disorders | Alzheimer's disease and other dementias; Parkinson's disease; Multiple sclerosis; Motor neuron disease; Other neurological disorders; Idiopathic developmental intellectual disability; Down syndrome; Neural tube defects |
| 11 | Sickle cell disorders | Sickle cell disorders |

\* we excluded latent tuberculosis and adjusted asthma to better reflect BTS steps 4+.

**Table 2.** Low estimates of the number of individuals in millions (% of total population) at increased risk of severe COVID-19 illness by age, number of conditions, region and age threshold.

|  | Africa | Asia | Europe | Latin America and the Caribbean | Northern America | Oceania | Global |
| --- | --- | --- | --- | --- | --- | --- | --- |
| **No conditions** |  |  |  |  |  |  |  |
| <15 years | 467 (40) | 948 (23) | 109 (16) | 137 (24) | 59 (18) | 9 (23) | **1729 (25)** |
| 15-49 years | 508 (43) | 1910 (46) | 272 (40) | 276 (48) | 139 (42) | 17 (43) | **3122 (45)** |
| 50-54 years | 27 (2) | 195 (5) | 36 (5) | 24 (4) | 15 (5) | 2 (4) | **299 (4)** |
| 55-59 years | 20 (2) | 152 (4) | 34 (5) | 20 (3) | 15 (4) | 2 (4) | **242 (3)** |
| 60-64 years | 14 (1) | 113 (3) | 28 (4) | 15 (3) | 12 (4) | 1 (3) | **184 (3)** |
| 65-69 years | 9 (1) | 87 (2) | 22 (3) | 11 (2) | 9 (3) | 1 (2) | **139 (2)** |
| 70+ years | 11 (1) | 109 (3) | 40 (6) | 15 (3) | 15 (5) | 2 (4) | **192 (3)** |
| All ages | 1056 (90) | 3513 (85) | 542 (79) | 499 (86) | 265 (80) | 32 (85) | **5906 (85)** |
| **One condition only** |  |  |  |  |  |  |  |
| <15 years | 8 (1) | 21 (1) | 1 (0) | 1 (0) | 0 (0) | 0 (0) | **31 (0)** |
| 15-49 years | 59 (5) | 215 (5) | 29 (4) | 26 (5) | 11 (3) | 2 (4) | **342 (5)** |
| 50-54 years | 9 (1) | 52 (1) | 9 (1) | 7 (1) | 4 (1) | 0 (1) | **81 (1)** |
| 55-59 years | 8 (1) | 50 (1) | 11 (2) | 7 (1) | 6 (2) | 0 (1) | **82 (1)** |
| 60-64 years | 7 (1) | 45 (1) | 12 (2) | 6 (1) | 6 (2) | 0 (1) | **77 (1)** |
| 65-69 years | 6 (0) | 41 (1) | 12 (2) | 5 (1) | 6 (2) | 0 (1) | **71 (1)** |
| 70+ years | 8 (1) | 67 (2) | 30 (4) | 11 (2) | 13 (4) | 1 (3) | **130 (2)** |
| All ages | 104 (9) | 491 (12) | 105 (15) | 63 (11) | 47 (14) | 4 (12) | **814 (12)** |
| **Multiple (two or more) conditions** |  |  |  |  |  |  |  |
| <15 years | 0 (0) | 0 (0) | 0 (0) | 0 (0) | 0 (0) | 0 (0) | **0 (0)** |
| 15-49 years | 5 (0) | 20 (0) | 3 (0) | 3 (0) | 1 (0) | 0 (0) | **32 (0)** |
| 50-54 years | 2 (0) | 11 (0) | 2 (0) | 1 (0) | 1 (0) | 0 (0) | **17 (0)** |
| 55-59 years | 2 (0) | 14 (0) | 3 (0) | 2 (0) | 2 (0) | 0 (0) | **22 (0)** |
| 60-64 years | 2 (0) | 16 (0) | 4 (1) | 2 (0) | 2 (1) | 0 (0) | **27 (0)** |
| 65-69 years | 2 (0) | 18 (0) | 5 (1) | 2 (0) | 3 (1) | 0 (0) | **31 (0)** |
| 70+ years | 5 (0) | 45 (1) | 22 (3) | 7 (1) | 9 (3) | 1 (2) | **89 (1)** |
| All ages | 19 (2) | 124 (3) | 39 (6) | 18 (3) | 18 (5) | 1 (4) | **218 (3)** |
| **At increased risk of severe COVID-19 illness** |  |  |  |  |  |  |  |
| At least one condition | 123 (10) | 614 (15) | 144 (21) | 81 (14) | 64 (20) | 6 (15) | **1032 (15)** |
| ...plus age 50+ years | 204 (17) | 1269 (31) | 305 (44) | 166 (29) | 131 (40) | 13 (33) | **2088 (30)** |
| ...plus age 55+ years | 178 (15) | 1074 (26) | 268 (39) | 142 (24) | 116 (35) | 11 (29) | **1789 (26)** |
| ...plus age 60+ years | 157 (13) | 923 (22) | 234 (34) | 122 (21) | 101 (31) | 10 (25) | **1547 (22)** |
| ...plus age 65+ years | 136 (12) | 722 (17) | 187 (27) | 97 (17) | 83 (25) | 7 (19) | **1233 (18)** |
| ...plus age 70+ years | 131 (11) | 681 (17) | 175 (25) | 92 (16) | 77 (24) | 7 (18) | **1163 (17)** |

**Table 3.** High estimates of the number of individuals in millions (% of total population) at increased risk of severe COVID-19 illness by age, number of conditions, region and choice of age threshold.

|  | Africa | Asia | Europe | Latin America and the Caribbean | Northern America | Oceania | Global |
| --- | --- | --- | --- | --- | --- | --- | --- |
| **No conditions** |  |  |  |  |  |  |  |
| <15 years | 577 (38) | 1140 (22) | 126 (16) | 169 (23) | 72 (18) | 10 (23) | **2093 (24)** |
| 15-49 years | 560 (37) | 2059 (40) | 279 (34) | 308 (43) | 156 (38) | 17 (38) | **3378 (39)** |
| 50-54 years | 23 (2) | 175 (3) | 31 (4) | 22 (3) | 14 (3) | 1 (3) | **267 (3)** |
| 55-59 years | 16 (1) | 126 (2) | 26 (3) | 16 (2) | 12 (3) | 1 (3) | **197 (2)** |
| 60-64 years | 10 (1) | 83 (2) | 18 (2) | 11 (1) | 8 (2) | 1 (2) | **131 (2)** |
| 65-69 years | 5 (0) | 56 (1) | 12 (1) | 6 (1) | 5 (1) | 1 (1) | **85 (1)** |
| 70+ years | 4 (0) | 48 (1) | 12 (1) | 6 (1) | 4 (1) | 1 (1) | **73 (1)** |
| All ages | 1193 (80) | 3686 (72) | 504 (62) | 538 (74) | 271 (66) | 32 (71) | **6224 (72)** |
| **One condition only** |  |  |  |  |  |  |  |
| <15 years | 26 (2) | 61 (1) | 4 (0) | 4 (1) | 2 (0) | 0 (1) | **96 (1)** |
| 15-49 years | 139 (9) | 509 (10) | 66 (8) | 61 (8) | 27 (7) | 4 (8) | **807 (9)** |
| 50-54 years | 18 (1) | 108 (2) | 18 (2) | 14 (2) | 9 (2) | 1 (2) | **167 (2)** |
| 55-59 years | 16 (1) | 100 (2) | 22 (3) | 13 (2) | 11 (3) | 1 (2) | **162 (2)** |
| 60-64 years | 13 (1) | 88 (2) | 23 (3) | 12 (2) | 12 (3) | 1 (2) | **148 (2)** |
| 65-69 years | 10 (1) | 77 (1) | 21 (3) | 10 (1) | 11 (3) | 1 (2) | **130 (2)** |
| 70+ years | 13 (1) | 107 (2) | 43 (5) | 17 (2) | 20 (5) | 2 (4) | **201 (2)** |
| All ages | 236 (16) | 1050 (20) | 197 (24) | 131 (18) | 90 (22) | 9 (20) | **1712 (20)** |
| **Multiple (two or more) conditions** |  |  |  |  |  |  |  |
| <15 years | 3 (0) | 8 (0) | 1 (0) | 1 (0) | 0 (0) | 0 (0) | **13 (0)** |
| 15-49 years | 26 (2) | 102 (2) | 14 (2) | 12 (2) | 6 (1) | 1 (2) | **161 (2)** |
| 50-54 years | 6 (0) | 38 (1) | 6 (1) | 5 (1) | 3 (1) | 0 (1) | **58 (1)** |
| 55-59 years | 7 (0) | 42 (1) | 9 (1) | 6 (1) | 4 (1) | 0 (1) | **68 (1)** |
| 60-64 years | 7 (0) | 45 (1) | 12 (1) | 6 (1) | 6 (1) | 0 (1) | **75 (1)** |
| 65-69 years | 7 (0) | 48 (1) | 13 (2) | 6 (1) | 7 (2) | 0 (1) | **82 (1)** |
| 70+ years | 14 (1) | 118 (2) | 52 (6) | 19 (3) | 23 (6) | 2 (4) | **228 (3)** |
| All ages | 69 (5) | 400 (8) | 107 (13) | 55 (8) | 49 (12) | 4 (9) | **685 (8)** |
| **At increased risk of severe COVID-19 illness** |  |  |  |  |  |  |  |
| At least one condition | 305 (20) | 1450 (28) | 304 (38) | 186 (26) | 139 (34) | 13 (29) | **2398 (28)** |
| ...plus age 50+ years | 362 (24) | 1937 (38) | 404 (50) | 247 (34) | 182 (44) | 18 (39) | **3150 (37)** |
| ...plus age 55+ years | 339 (23) | 1762 (34) | 373 (46) | 225 (31) | 168 (41) | 16 (36) | **2884 (33)** |
| ...plus age 60+ years | 323 (22) | 1637 (32) | 346 (43) | 209 (29) | 156 (38) | 15 (33) | **2686 (31)** |
| ...plus age 65+ years | 329 (22) | 1634 (32) | 369 (46) | 213 (29) | 169 (41) | 16 (35) | **2729 (32)** |
| ...plus age 70+ years | 319 (21) | 1557 (30) | 348 (43) | 203 (28) | 158 (39) | 15 (33) | **2599 (30)** |

